# Enabling a Learning Public Health System: Enhanced Surveillance of HIV and Other Sexually Transmitted Infections

**DOI:** 10.1101/2024.04.10.24305612

**Authors:** Harry Reyes Nieva, Jason Zucker, Emma Tucker, Delivette Castor, Michael T. Yin, Peter Gordon, Noémie Elhadad

## Abstract

Sexually transmitted infections (STIs) continue to pose a substantial public health challenge in the United States (US). Surveillance, a cornerstone of disease control and prevention, can be strengthened to promote more timely, efficient, and equitable practices by incorporating health information exchange (HIE) and other large-scale health data sources into reporting. New York City patient-level electronic health record data between January 1, 2018 and June 30, 2023 were obtained from Healthix, the largest US public HIE. Healthix data were linked to neighborhood-level information from the American Community Survey. In this cross-sectional study, we compared patients who received a test or tested positive for chlamydia, gonorrhea, and/or HIV with patients who were untested or tested negative, respectively, using generalized estimating equations with logit function and robust standard errors. Among 1,519,121 tests performed for chlamydia, 1,574,772 for gonorrhea, and 1,200,560 for HIV, 2%, 0.6% and 0.3% were positive for chlamydia, gonorrhea, and HIV, respectively. Chlamydia and gonorrhea co-occurred in 1,854 cases (7% of chlamydia and 21% of gonorrhea total cases). Testing behavior was often incongruent with geographic and sociodemographic patterns of positive cases. For example, people living in areas with the highest levels of poverty were less likely to test for gonorrhea but almost twice as likely to test positive compared to those in low poverty areas. Regional HIE enabled review of testing and cases using granular and complementary data not typically available given existing reporting practices. Enhanced surveillance spotlights potential incongruencies between testing patterns and STI risk in certain populations, signaling potential under- and over-testing. These and future insights derived from HIE data may be used to continuously inform public health practice and drive further improvements in provision and evaluation of services and programs.

## INTRODUCTION

Sexually transmitted infections (STIs) continue to pose a substantial public health challenge. The Centers for Disease Control and Prevention (CDC) estimate there are about 67.6 million prevalent and 26.2 million incident cases of STIs in the United States.^1^ These statistics largely result from steady increases in rates of common nationally notifiable STIs such as chlamydia and gonorrhea over the last decade. During this same period, longstanding disparities in human immunodeficiency virus (HIV) and other STIs persist unabated among historically minoritized populations. For example, people who identify as Black or African American comprise 40% of new HIV diagnoses despite representing only 14% of the population.^2^

Traditionally, prevalence and incidence estimates are based on STI surveillance systems that rely heavily on direct notification from healthcare providers and laboratories. Due to this, only positive cases are typically reported to local health agencies as part of standard clinical practice. Lack of reporting on testing (e.g., number conducted) results in lack of information on positivity rates, limiting understanding of differences in population-level risk for public health services and programs. In addition, delays in case reporting and long lag time required for data aggregation and preparation of surveillance reports (often at least 9 months following the review period) substantially hinder real-time response to potential outbreaks.^3^ Furthermore, fulfilling this vital mandate of public health agencies is often hampered by an ailing and highly fragmented data infrastructure precluding reliable, timely, and accurate data.^4^

Numerous agencies have prioritized addressing health inequity through enhanced analytical capacity.^3,5^ Surveillance may be strengthened by incorporating rich, complementary data sources such as readily available public datasets (e.g., national survey data) and regional health information exchanges (HIEs).^6^ HIEs share electronic health record (EHR) data across multiple health systems and EHR platforms within a defined geographic area, allowing for aggregation of data in a fashion that closely approximates regional surveillance performed by departments of public health. Utilization of HIE data also affords the added advantage of granular, timely, and comprehensive clinical and demographic information typically unavailable using existing reporting practices.^7,8^ Prior studies involving HIE use have been associated with both cost and time savings on demographic and treatment requests made by public health staff,^9^ decrease in unnecessary diagnostic testing in emergency departments,^10^ improved continuity of care and patient outcomes,^11–14^ and a reduction in racial and ethnic disparities.^15^ Large publicly available datasets offer additional context when data linkages are made based on common geography, an important dimension of social determinants of health (SDOH).^16,17^

The objective of this study was to demonstrate how comprehensive, near real-time HIE data and a complementary, large public data source might be leveraged to promote timely and equitable surveillance of HIV and other STIs in support of existing public health reporting. Specifically, we sought to: 1) quantify the number of laboratory tests conducted and percent of tests that resulted in confirmed positive cases of chlamydia, gonorrhea, and HIV; 2) examine trends in concurrent testing for and co-occurring cases of chlamydia, gonorrhea, and HIV; and 3) characterize population-level differences between testing and positivity rates for chlamydia, gonorrhea, and HIV along geographic and sociodemographic dimensions.

## METHODS

### Study design, setting, and population

We performed a retrospective observational cross-sectional study of adults residing and receiving care in New York City (NYC) between January 2018 and June 2023. We examined testing and test results for chlamydia, gonorrhea, and HIV. Laboratory testing included nucleic acid amplification for chlamydia and gonorrhea and serology testing for HIV. We compared patients who received a test or had a positive test for a given STI with patients who were untested or had a negative test, respectively. This study was approved by the institutional review board of Columbia University Irving Medical Center (Protocol AAAT1774) and reporting follows the Strengthening the Reporting of Observational Studies in Epidemiology (STROBE) guideline.

### Data

Our primary data source was Healthix, the largest public health information exchange (HIE) in the United States, which serves the NYC metropolitan area and Long Island, New York. Funded by the New York State Department of Health, Healthix collects EHR and administrative data from more than 8,000 healthcare facilities for over 20 million patients. Healthix uses a proprietary probabilistic matching system paired with human intelligence to accurately match data collected for the same patient across multiple care sites.^18^ Healthix provided a limited dataset via secure file transfer protocol (SFTP) with patient information including demographics, diagnoses, encounters, procedures, lab results, and medications. For each patient in this extract, we requested all chlamydia, gonorrhea, HIV screening tests available. All data files were stored on a secure Linux server compliant with Health Insurance Portability and Accountability Act (HIPAA) standards.

We used the Observational Health Data Sciences and Informatics (OHDSI) Observational Medical Outcomes Partnership (OMOP) Common Data Model (CDM) to standardize EHR and laboratory testing center data from different health systems and facilities.^19^ Source values for patient sex may have represented sex assigned at birth or the administrative sex used for billing purposes (which may reflect gender identity). Race and ethnicity were also often conflated in the source data, in particular with respect to persons identifying as Hispanic, Latina, Latino, Latine, or Latinx. For brevity, we use “Hispanic or Latino.” HIV cases were classified using the Recommended Laboratory HIV Testing Algorithm for Serum or Plasma Specimens.^20^ Laboratory tests for previously known positive cases and HIV viral load monitoring were removed from the dataset.

For each patient, we derived United Hospital Fund (UHF) neighborhood designations based on grouping of contiguous residential ZIP Code Tabulation Areas (**Table S1**).^21^ The NYC Department of Health and Mental Hygiene (DOHMH) commonly uses UHF neighborhood designations in STI surveillance reports. We also converted patient residential ZIP code to ZIP Code Tabulation Area (ZCTA) to identify area-based poverty level (ABPL) based on 2020 American Community Survey (ACS) 5-Year Estimates using the Census Bureau’s Application Programming Interface (API).^22^ ABPL, defined as the percentage of people earning below the Federal Poverty Threshold (FPT) within a ZCTA, is a standard measure of socioeconomic status (SES) used by the NYC DOHMH to describe and monitor disparities in public health data analyses, including STI reporting.^23,24^ In accordance with NYC DOHMH standard cut-points, we categorized neighborhood-level poverty as low, medium, high, and very high (<10%, 10% to <20%, 20% to <30%, 30% to 100% of residents in ZCTA living below the FPT, respectively). Patient ages were grouped into categories based on intervals used in recent NYC DOHMH reports (e.g., 20-24, 55-59, 65+).^25^

### Outcomes and Variables

The primary outcomes were 1) occurrence of a laboratory test for chlamydia, gonorrhea, or HIV and 2) determination of a positive test result. For each STI of interest, we examined trends overall and differences based on factors such as patient sex, race, ethnicity, age, residential neighborhood, borough, and ABPL.

### Statistical Analysis and Data Visualization

We computed descriptive statistics, including assessment of concurrent testing and co-occurring positive cases, using counts and percentages. We compared proportions using the *χ*^2^ test and means using the Student’s t-test. To assess neighborhood-level clustering of rates, we computed Moran’s I statistics. To estimate the odds of testing for a given STI or obtaining a positive test result, we fit unadjusted and adjusted logistic regression models using generalized estimating equations with an exchangeable correlation structure to account for clustering of multiple tests for a given patient. We computed unadjusted and adjusted odds ratios and 95% confidence intervals (CIs) using robust standard errors. Models examined patient sex, race, ethnicity, age, borough of residence, and ABPL. To compute absolute difference between case and test percentages, we divided the number of tests or cases in each UHF neighborhood by citywide totals, then subtracted the percentage of tests performed from the percentage of cases identified (e.g., a positive difference indicates more citywide cases than tests in the neighborhood). We conducted all statistical analyses with p<0.05 indicating statistical significance and generated chloropleth maps using the R programming environment, version 4.1.1 (R Foundation for Statistical Computing, Vienna, Austria).

## RESULTS

The HIE dataset comprised 4,767,322 patients with a mean age of 46 (standard deviation [SD], 18) years; 61% were identified as female and 38% as male (**Table 1**). Among patients, 14% were identified as Asian or Pacific Islander, 17% as Black or African American, 18% as Hispanic or Latino, 21% as White; 11% were labeled as “Other” in source data, and 19% were of unknown race. Generally, one to three percent of patients resided in each of the 42 neighborhoods in NYC (**Fig. 1**). Approximately 66% of patients lived in areas of low or medium ABPL and 33% in areas of high or very high ABPL (**Table 1**). Sociodemographic differences in the proportion of persons tested compared to those untested and persons with positive test results versus those with negative results were statistically significant (p<0.001 for all comparisons).

**Figure 1:**
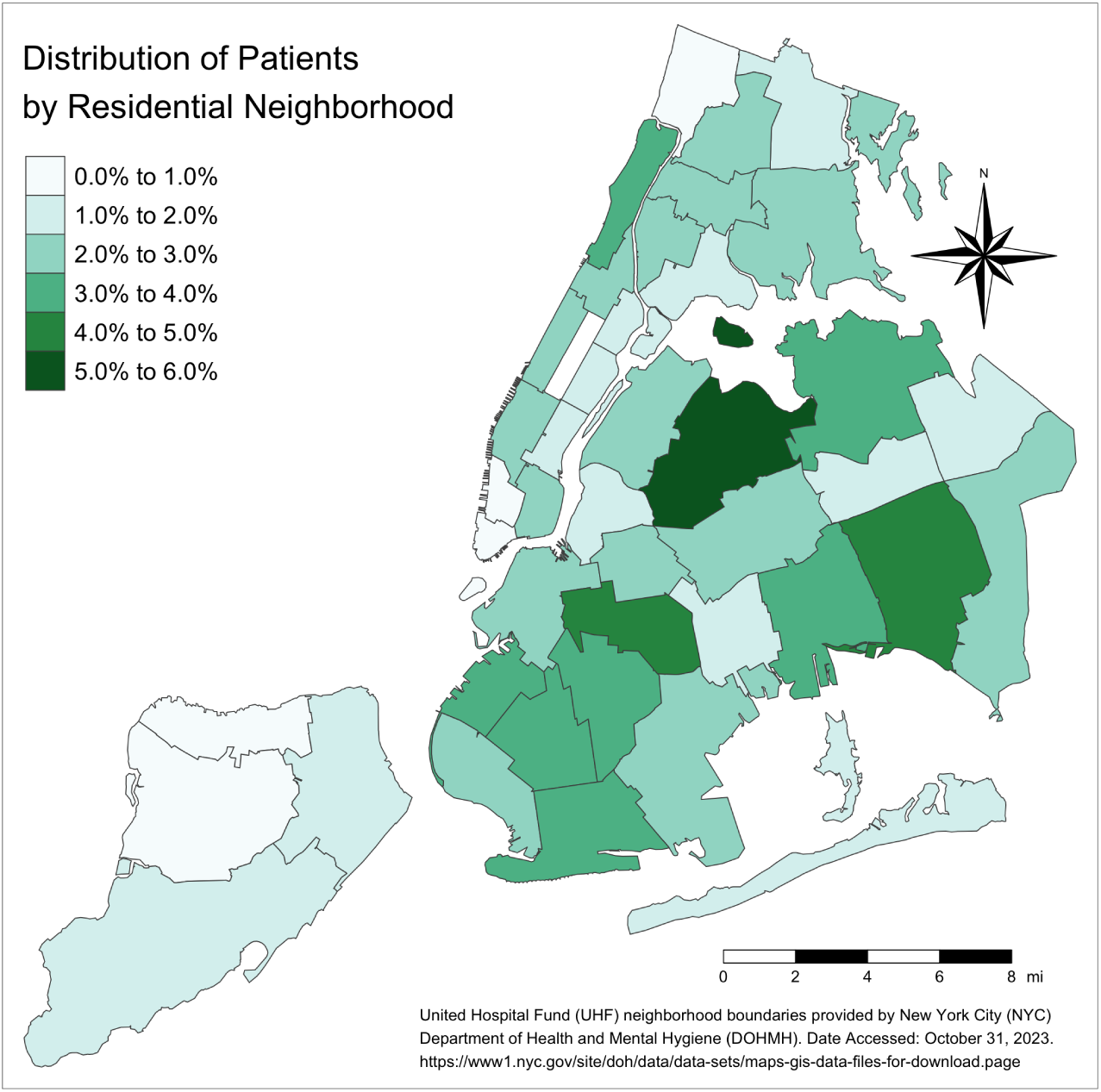
Geographic distribution of patients in the cohort by residential neighborhood.

**Table 1:** Cohort characteristics.

| Characteristics | Total<br>N=4,767,322 patients | Chlamydia |  | Gonorrhea |  | HIV |  |
| --- | --- | --- | --- | --- | --- | --- | --- |
|  |  | Testing<br>N=1,519,121 tests | Positive Result<br>N=28,251 tests | Testing<br>N=1,574,772 tests | Positive Result<br>N=8,873 tests | Testing<br>N=1,200,560 tests | Positive Result<br>N=3,374 tests |
|  |  | mean (SD) |  |  |  |  |  |
| Age, years | 46 (18) | 36 (13) | 27 (9) | 36 (13) | 29 (10) | 39 (15) | 42 (14) |
|  |  | N (%) |  |  |  |  |  |
| **Sex<sup>a</sup>** |  |  |  |  |  |  |  |
| Female | 2,922,650 (61) | 1,075,218 (71) | 19,299 (67) | 1,112,482 (71) | 3,417 (39) | 751,845 (63) | 852 (25) |
| Male | 1,806,333 (38) | 438,526 (29) | 9,390 (33) | 456,964 (29) | 5,424 (61) | 442,764 (37) | 2,490 (74) |
| Unknown | 38,339 (1) | 5,377 (0) | 59 (0) | 5,326 (0) | 32 (0) | 5,951 (1) | 32 (1) |
| **Race or Ethnicity<sup>b</sup>** |  |  |  |  |  |  |  |
| Asian or Pacific Islander | 674,314 (14) | 165,610 (11) | 1,647 (6) | 171,496 (11) | 302 (3) | 146,649 (12) | 86 (3) |
| Black or African American | 807,649 (17) | 289,846 (19) | 8,678 (30) | 302,873 (19) | 3,682 (41) | 229,949 (19) | 1,235 (37) |
| Hispanic or Latino | 850,799 (18) | 342,745 (23) | 7,576 (26) | 355,666 (23) | 1,596 (18) | 254,122 (21) | 739 (22) |
| White | 1,002,214 (21) | 263,821 (17) | 2,583 (9) | 276,510 (18) | 1,014 (11) | 214,989 (18) | 392 (12) |
| Other race<sup>c</sup> | 525,063 (11) | 164,863 (11) | 3,139 (11) | 169,584 (11) | 886 (10) | 134,870 (11) | 352 (10) |
| Unknown | 907,283 (19) | 292,236 (19) | 5,125 (18) | 298,643 (19) | 1,393 (16) | 219,981 (18) | 570 (17) |
| **Borough of Residence** |  |  |  |  |  |  |  |
| Bronx | 662,357 (14) | 271,499 (18) | 6,421 (22) | 272,804 (17) | 1,921 (22) | 194,505 (16) | 913 (27) |
| Brooklyn | 1,492,574 (31) | 485,206 (32) | 8,538 (30) | 501,624 (32) | 2,841 (32) | 392,844 (33) | 1,054 (31) |
| Manhattan | 1,457,898 (31) | 418,765 (28) | 4,854 (17) | 440,551 (28) | 1,657 (19) | 344,493 (29) | 610 (18) |
| Queens | 888,620 (19) | 275,909 (18) | 7,734 (27) | 287,022 (18) | 2,034 (23) | 217,985 (18) | 692 (21) |
| Staten Island | 265,873 (6) | 67,742 (5) | 1,201 (4) | 72,771 (5) | 420 (5) | 50,733 (4) | 105 (3) |
| **Area-based Poverty Level<sup>d</sup>** |  |  |  |  |  |  |  |
| Low (<10% below FPT) | 919,691 (19) | 247,611 (16) | 3,972 (14) | 264,626 (17) | 947 (11) | 202,253 (17) | 319 (10) |
| Medium (10% to <20%) | 2,250,976 (47) | 676,764 (45) | 11,593 (40) | 701,128 (45) | 3,497 (39) | 551,405 (46) | 1,443 (43) |
| High (20% to <30%) | 959,814 (20) | 340,222 (22) | 6,896 (24) | 350,817 (22) | 2,284 (26) | 258,018 (21) | 766 (23) |
| Very high (30% to 100%) | 635,296 (13) | 253,967 (17) | 6,269 (22) | 257,622 (16) | 2,141 (24) | 188,455 (16) | 843 (25) |
| Unknown | 1,545 (0) | 557 (0) | 18 (0) | 579 (0) | 4 (0) | 429 (0) | 3 (0) |

Abbreviations: ABPL, Area-based Poverty Level. CDM, Common Data Model. FPT, federal poverty threshold. HIE, health information exchange. HIV, human immunodeficiency virus. OMOP, Observational Medical Outcomes Partnership. UHF, United Hospital Fund. <sup>a</sup>The OMOP CDM uses the term “gender” while source data were based on sex; source values may represent sex assigned at birth and/or administrative sex used for billing purposes (which may reflect gender identity, not sex assigned at birth). <sup>b</sup>All categories other than “Hispanic or Latino” are exclusively non-Hispanic or Latino (e.g., non-Hispanic White). <sup>c</sup>“Other Race” category is derived directly from reporting of “Other”, “Other race”, or similar in the source data. <sup>d</sup>ABPL categorized as low (<10% of residents in ZIP code tabulation area living below the FPT), medium (10% to <20%), high (20 to <30%), and very high (30 to 100%).

### Diagnostic Testing and Positivity Rates

During the review period, patients received 1,519,121 tests for chlamydia, 1,574,772 tests for gonorrhea, and 1,200,560 for HIV (**Fig. 2**). Among tests conducted, 28,251 (2%) were positive for chlamydia (1,860 positive per 100,000 tests), 8,873 (0.6%) for gonorrhea (563 positive per 100,000 tests), and 3,374 (0.3%) for HIV (281 positive per 100,000 tests).

**Figure 2:**
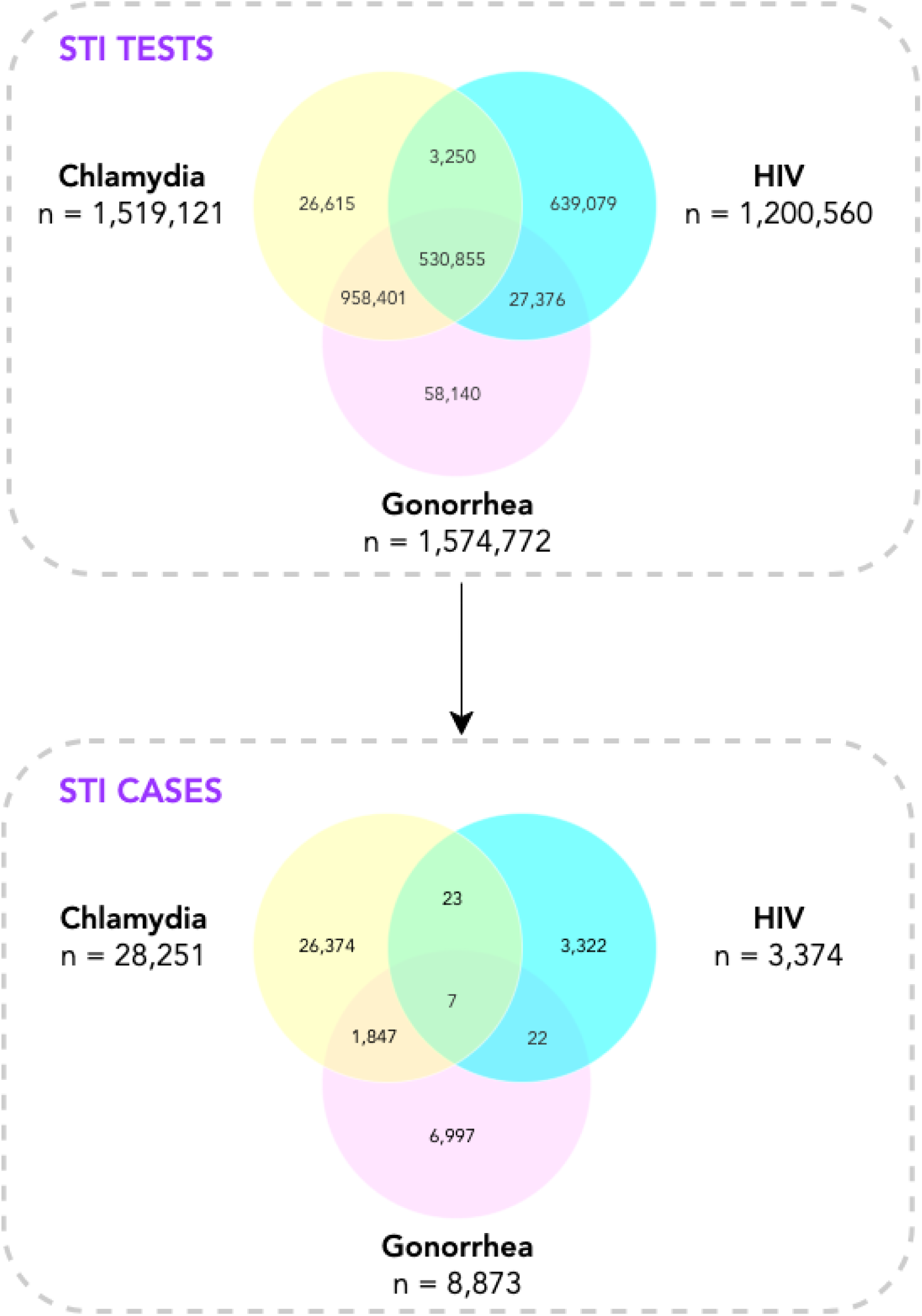
Concurrent laboratory testing and co-occurring cases of chlamydia, gonorrhea, and HIV.

### Concurrent Testing and Co-infection

Chlamydia and gonorrhea testing were largely concurrent (1,489,256 shared tests) comprising 98% of chlamydia tests and 95% of gonorrhea tests (**Fig. 2**). HIV testing in combination with testing for both chlamydia and gonorrhea (n=530,855) represented 44% of HIV tests (35% and 34% of chlamydia and gonorrhea tests, respectively). Dual testing for HIV and gonorrhea (n=27,376) was more common than dual testing for HIV and chlamydia (n=3,250). Single testing for HIV comprised 53% of all HIV testing. Testing was positive for both chlamydia and gonorrhea in 1,854 tests (7% of chlamydia and 21% of gonorrhea total cases). Results were positive for HIV and chlamydia in 23 tests (0.7% of HIV and 0.1% of chlamydia total cases), positive for both HIV and gonorrhea in 22 tests (0.6% of HIV and 0.3% of gonorrhea total cases), and positive for all three STIs in seven tests. One-third of positive chlamydia cases (34%) had an HIV test performed during the same visit (9,659 of 28,251 total cases). Nearly one-fifth (22%) of positive gonorrhea cases had an HIV test performed during the same visit (1,954 of 8,873 total cases).

### Testing and Positivity by Population

We identified population-level trends concerning testing behavior and positive cases (**Fig. 3**, **Tables S2-3**). Men were nearly half as likely to test for chlamydia (adjusted odds ratio [aOR]:0.62, 95% CI:0.62-0.63) or gonorrhea (aOR:0.63, 95%CI:0.63-0.63) and more likely to test for HIV as women (aOR:1.16, 95%CI:1.15-1.17). In contrast, men were slightly more likely to receive a positive result for chlamydia (aOR:1.09, 95%CI:1.05-1.12), three times as likely to test positive for gonorrhea (aOR:3.28, 95%CI:3.11-3.45), and five times as likely to receive a positive test result for HIV (aOR:5.18, 95%CI:4.79-5.61).

**Figure 3:**
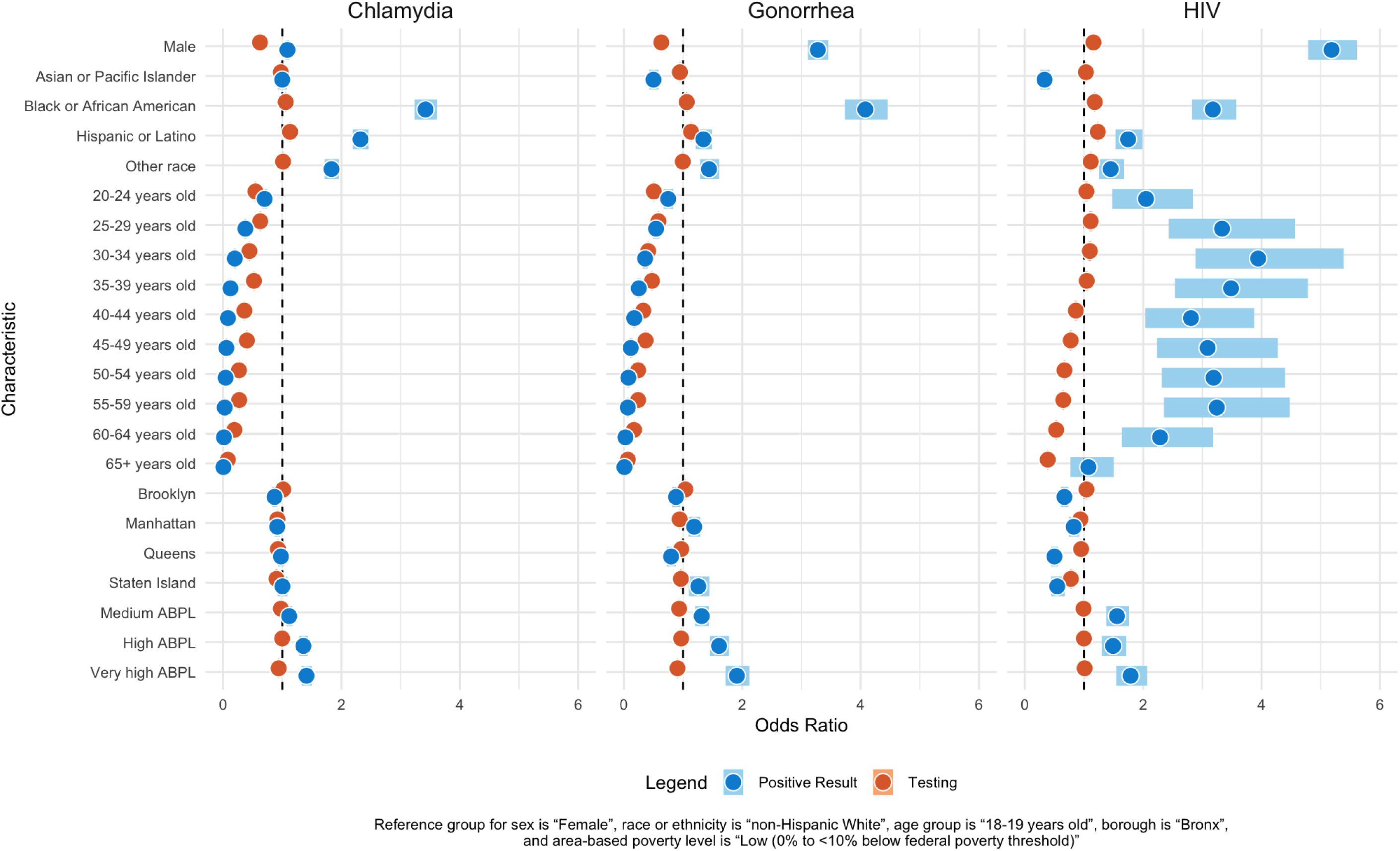
Forest plots of adjusted odds ratios for laboratory testing and positive test results by sexually transmitted infection and patient characteristics.

Patients who were Asian or Pacific Islander were nearly as likely to test for gonorrhea or HIV as those who were White (aOR:0.95, 95%CI:0.94-0.95 and aOR:1.03, 95%CI:1.02-1.04, respectively) but far less likely to test positive (aOR:0.50, 95%CI:0.43-0.58 and aOR:0.33, 95%CI:0.26-0.42, respectively). Participants who were Black or African American were nearly as likely to get tested for chlamydia, gonorrhea, or HIV but their odds of receiving a positive result were three to four times as high as their odds of taking a test (e.g., aOR:4.08, 95%CI:3.73-4.46 for positive gonorrhea test compared to aOR:1.06, 95%CI:1.05-1.07 for gonorrhea testing). A similar but smaller difference was also notable in chlamydia testing among patients identified as Hispanic or Latino (aOR:1.13, 95%CI:1.12-1.14 for positive result versus aOR:2.32, 95%CI:2.19-2.46 for testing).

Notably, as age increased, odds of testing and odds of receiving a positive test result for chlamydia and gonorrhea decreased. Likelihood of testing for HIV decreased with age as well, though odds of testing positive did not follow this same trend. Compared to patients in the 18-19 years old category, patients who were 30-34 years old had the highest odds of testing positive (aOR:3.94, 95%CI:2.88-5.39) followed by patients who were 35-39 years old (aOR:3.49, 95%CI:2.54-4.78).

Differences in odds of testing and positivity were also found among patients given the poverty level of the area in which they lived. Compared to patients living in areas with low levels of poverty, those residing in areas with high levels of poverty were about as likely to get tested but were more likely to test positive for chlamydia (aOR:1.36, 95%CI:1.29-1.43), gonorrhea (aOR:1.61, 95%CI:1.45-1.77), and HIV (aOR:1.49, 95%CI:1.30-1.71). In contrast, patients living in areas with very high levels of poverty were less likely to test for either chlamydia (aOR:0.94, 95% CI:0.93-0.95) or gonorrhea (aOR:0.90, 95%CI:0.89-0.91) and more likely to test positive for all three STIs (aOR:1.41, 95%CI:1.33-1.50 for chlamydia; aOR:1.91, 95%CI:1.72-2.12 for gonorrhea; and aOR:1.79, 95%CI:1.55-2.07 for HIV).

### Distribution of STIs by Neighborhood

Testing and cases exhibited marked spatial heterogeneity (**Fig. 4**, **Table S4**) with positive spatial autocorrelation indicating statistically significant levels of clustering in both testing and cases (**Table S5**). The largest percentage of chlamydia, gonorrhea, and HIV tests were conducted in the West Queens neighborhood (5.9%, 5.8%, and 6.0%, respectively). By comparison, the highest percentage of positive cases for chlamydia, gonorrhea, and HIV (5.9%, 7.6%, and 7.8%, respectively) were in the Bedford Stuyvesant-Crown Heights neighborhood of Brooklyn (compared to 5.1%, 3.5%, and 3.9%, respectively, in West Queens).

**Figure 4:**
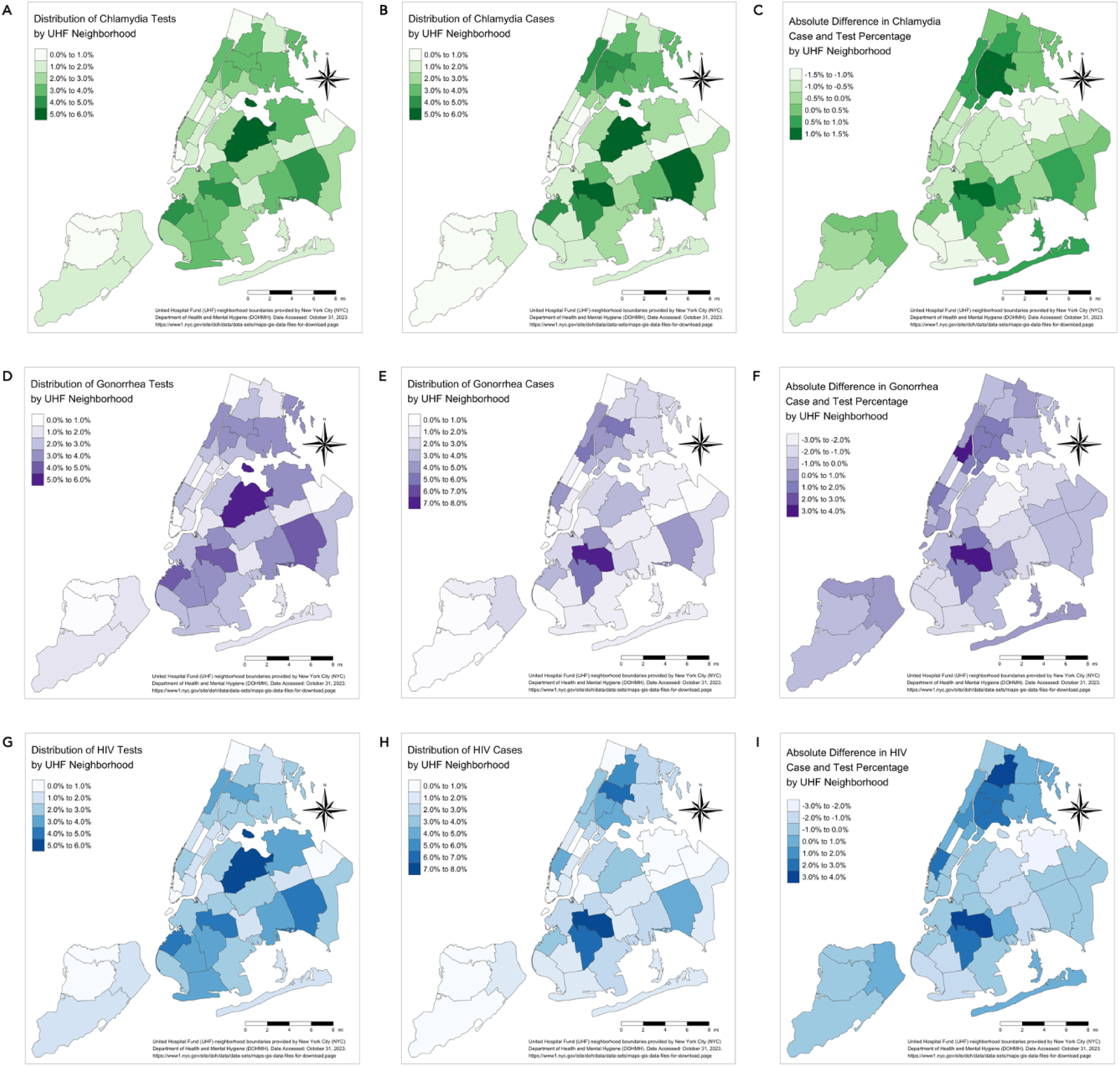
Geographic distribution of laboratory testing, positive cases, and absolute difference in case and test percentage of chlamydia, gonorrhea, and HIV in New York City by neighborhood.

The largest positive absolute differences between case and test percentages for chlamydia, gonorrhea, and HIV were consistently found in the Bedford Stuyvesant-Crown Heights neighborhood of Brooklyn (**Table S5**). Chlamydia comprised 1.4% more citywide positive cases than the percentage of citywide tests and 3.0% more gonorrhea and HIV citywide cases than tests. The largest negative absolute difference between case and test percentages for chlamydia was in the Borough Park neighborhood of Brooklyn (−1.4%). For gonorrhea, the largest negative absolute difference was in West Queens (−2.3%). Flushing, Queens had the largest negative absolute difference (−2.4%) for HIV.

## DISCUSSION

By leveraging data from a regional HIE with detailed, near real-time EHR data standardized across platforms and health systems, we established a robust, scalable data infrastructure to enhance STI surveillance, enable examination of population-level trends, and promote more effective and equitable testing and intervention. Our findings provide valuable information on laboratory-confirmed cases for chlamydia, gonorrhea, and HIV, in particular as they compare to underlying testing patterns and differ across populations and geographies.

The high burden of STIs, as evidenced by the large number of cases, underscores the importance of effective, timely surveillance and targeted interventions. Traditional surveillance heavily relies on direct notification from providers and laboratories, which limits understanding of population-level risk and the efficacy and equity of public services and programs. By incorporating data from HIEs and public datasets with neighborhood-level information relevant to SDOH, we were able to overcome these limitations and gain more comprehensive insights into STI testing and positivity rates.

Unsurprisingly, most patients had concurrent testing for chlamydia and gonorrhea as the most common nucleic acid amplification test performed is capable of identifying both diseases simultaneously. We did not, however, note similarly substantial concurrent testing for HIV in tandem with tests for chlamydia and gonorrhea. This underscores the need for implementing and strengthening integrated strategies that simultaneously screen for multiple STIs, as co-infections can have serious health consequences and contribute to ongoing transmission.^26–28^ By identifying the occurrence of concurrent testing and co-occurring infections, public health agencies can enhance their strategies for STI prevention and control by addressing missed opportunities for testing and implementing interventions to reduce repeated visits to seek care and the burden of multiple infections.

Enhanced surveillance also revealed notable disparities with respect to both distribution in and alignment between STI testing and positive cases among different socio-demographic groups. Consistent with previous studies, we found that members of certain populations were disproportionately affected by STIs. These findings highlight the urgent need for targeted interventions and strengthening of efforts to promote health equity.

Differences were also found when comparing testing and positivity rates based on patient sex, race, ethnicity, age, neighborhood, and socioeconomic status. Men were less likely to undergo testing for chlamydia and gonorrhea but had higher odds of receiving a positive result. This suggests potential missed opportunities for testing among men. This does not, however, lead us to recommend women receive less testing, as STIs are more commonly asymptomatic in women. Similarly, patients who were identified as Asian or Pacific Islander were almost as likely to test for HIV but far less likely to test positive compared to people who were identified as non-Hispanic white. People identified as Black, African American, Hispanic, or Latino were more likely to receive certain tests and test positive. Nonetheless, relative differences between testing and case rates signal possible undertesting. While STI testing generally decreased as age increased, odds of testing positive for HIV did not following this trend. Differences suggest missed opportunities to test. These findings highlight the complex interplay of socio-demographic factors in STI testing and positivity rates, emphasizing the importance of tailored interventions and culturally competent care. By utilizing HIE data, public health agencies may better identify and address disparities and tailor interventions to specific needs of different populations.

Geospatial analysis demonstrated marked heterogeneity in testing behavior and cases across neighborhoods and clustering of testing and cases. This suggests geography was a key factor among patterns of infection, diagnosis, and treatment. Unsurprisingly, this highlights the importance of the well-established practice of considering geographic in addition to demographic factors for STI prevention and control strategies. Our primary aim in presenting these expected results is to demonstrate the utility of HIE as a data source in characterizing spatial distribution of STIs to help identify high-risk areas, distinguish between sub-populations, and allocate resources more effectively and equitably (e.g., targeting interventions to specific areas via mobile testing sites).

It is widely reported that fragmented data collection systems at state and local levels have been underfunded for years and it has been estimated that modernizing non-federal public health reporting systems would require $8 billion per year for 10 years.^4^ In the era of big data, existing platforms like HIEs present opportunities to further empower public health professionals to conduct their work.^6^ This approach enables identification of emerging trends, prediction of outbreaks, and optimization of resource allocation for more targeted, effective population-based interventions. By continuously learning from real-world evidence, public health agencies may adapt strategies and responses to evolving STI dynamics more quickly, ultimately improving outcomes and reducing burden on communities.

Most notably, this data-driven approach aligns closely with a learning health system (LHS).^29^ The LHS framework can extend to public health systems which primarily operate at a population instead of an individual-service level.^30^ Indeed, a learning public health system (LPHS) might encompass multiple LHS and traditional health systems and routinely analyze large-scale datasets, extract insights regarding population health, and inform public health practice to enable continual improvement.

### Limitations

We acknowledge several limitations to our study in need of address to achieve an equitable LPHS. First, Healthix data did not capture all testing and cases within the NYC metropolitan area. Without a comprehensive list of non-Healthix facilities, we were unable to determine which sites were missing and whether certain clinic types or locations were over/underrepresented. Populations less well represented may have received testing at non-participating facilities. We also used residential ZIP code to perform geospatial analyses. While, our limited data set only allowed for spatial resolution to the ZIP code level, full residential addresses are available to the HIE. Thus, public health agencies may be able to access such information if needed to ensure more precise, targeted intervention. Notably, source data for patient sex and gender as well as race and ethnicity were often conflated in our dataset, which may introduce bias. Nearly 20% of patients were also missing data on race and/or ethnicity. An additional 10% were only assigned a value of “Other” in the source data. While these concerns are commonly found among EHR data, they remain a challenge if we are to support elucidation of disparities and offer actionable findings to inform intervention. In addition, patients on pre-exposure prophylaxis (PrEP) for HIV prevention often undergo regular STI testing. Although our modeling accounts for repeated measures from the same patient, our approach does not fully address how this may influence findings. Lastly, our analysis evaluated patterns related to chlamydia, gonorrhea, and HIV. Future work would be necessary to establish that other STIs (e.g., syphilis) reflect similar trends.

### Conclusion

Our study demonstrates the value of leveraging HIE and public data to enhance surveillance and inform interventions. By integrating detailed, near real-time data, public health agencies may gain a more nuanced understanding of testing patterns and positive cases, as well as identify disparities among different socio-demographic groups and geographic regions. This knowledge can guide development of targeted interventions, allocation of resources, and implementation of an equitable learning public health system. Moving forward, further research and collaboration between public health agencies, healthcare providers, and HIEs are needed to fully realize the potential of data-driven approaches in reducing the burden of STIs and improving community health outcomes.

## Acknowledgements

Preliminary data from this study were presented at the STI & HIV 2023 World Congress in Chicago, IL on July 25, 2023 and the American Medical Informatics Association 2023 Annual Symposium in New Orleans, LA on November 12, 2023.

## Contributor Statement

All authors had full access to the data in the study. They also take responsibility for the integrity of the data and accuracy of the analysis, and have approved the final manuscript.

*Concept and design:* Reyes Nieva, Elhadad, Zucker

*Acquisition, analysis, or interpretation of data:* All authors

*Drafting of the manuscript:* Reyes Nieva

*Critical revision of the manuscript for important intellectual content:* All authors

*Statistical analysis:* Reyes Nieva

*Obtained funding:* Elhadad, Zucker

*Administrative, technical, or material support:* Reyes Nieva, Tucker

*Supervision:* Elhadad, Zucker

## Data Availability Statement

The electronic health record and administrative data underlying this article are not available due to restrictions to preserve patient confidentiality.

## Supplemental Appendix

### Appendix A. Supplementary Figures

**Figure S1.**
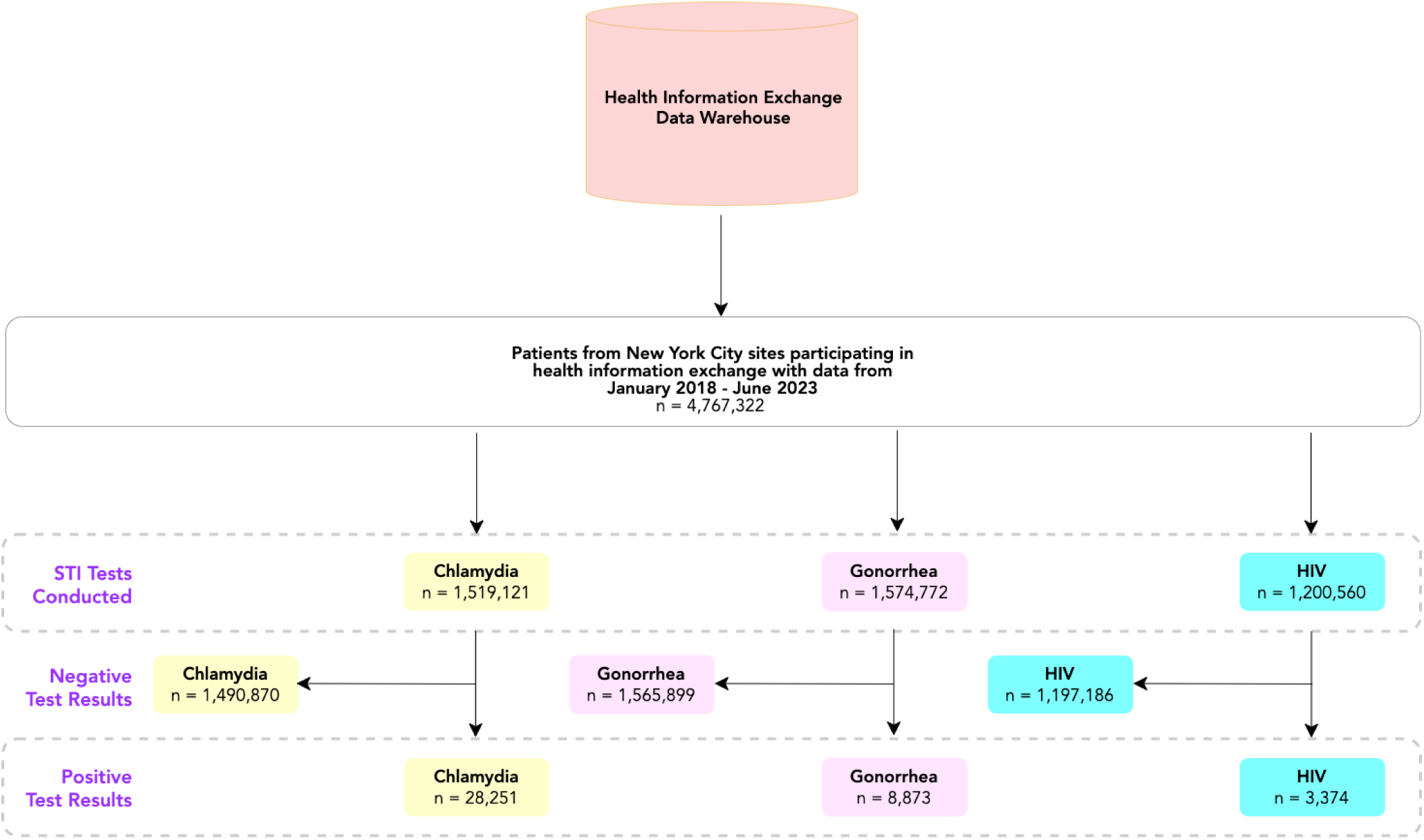
Strengthening the Reporting of Observational Studies in Epidemiology (STROBE) Statement flow chart.

### Appendix B. Supplementary Tables

**Table S1.**
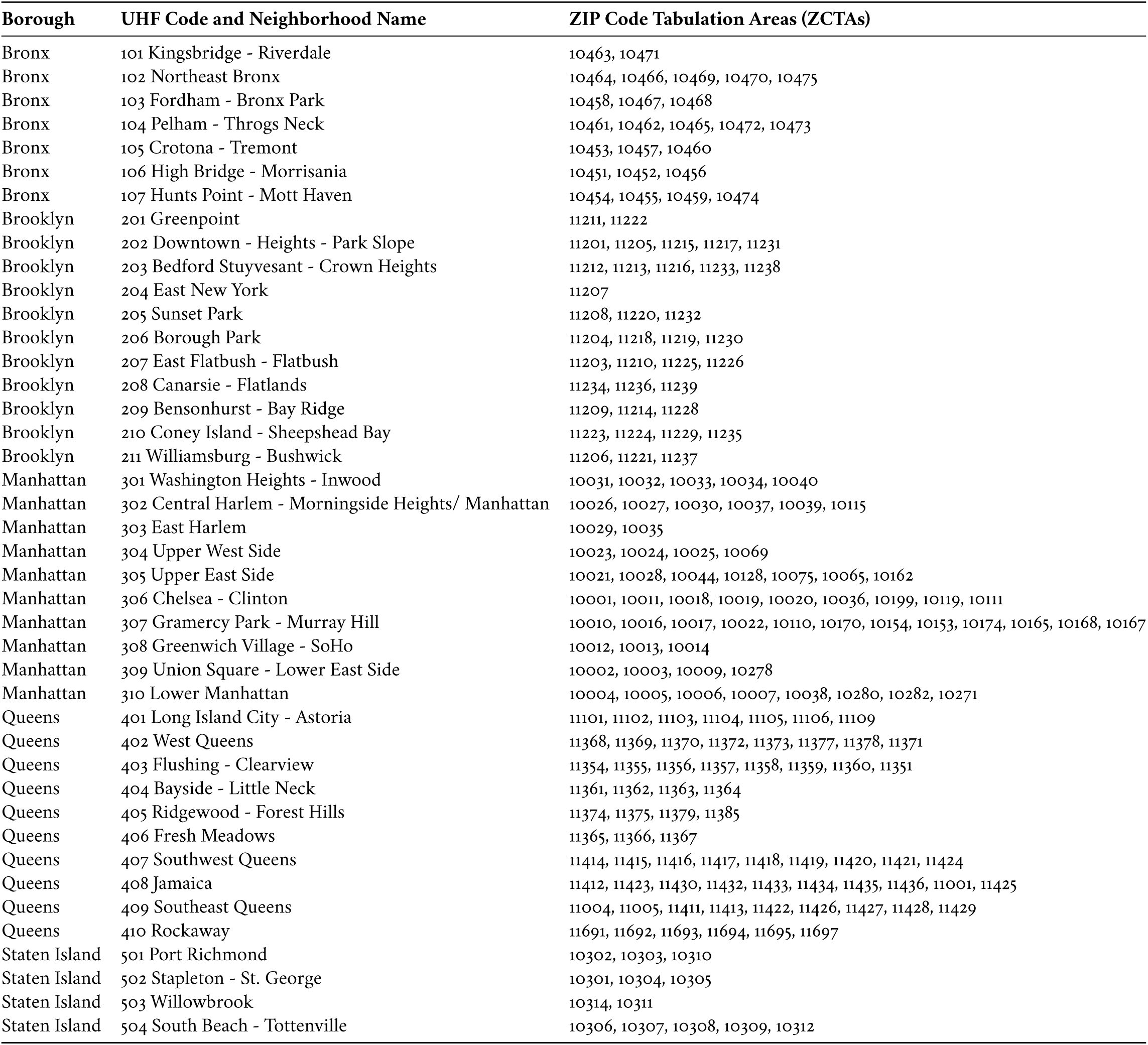
United Hospital Fund (UHF) neighborhood designation ZIP Code Tabulation Area (ZCTA) index.

**Table S2.** Adjusted odds ratios for laboratory testing and positive test results by sexually transmitted infection and patient characteristics.

| Characteristics | Chlamydia |  |  |  | Gonorrhea |  |  |  | HIV |  |  |  |
| --- | --- | --- | --- | --- | --- | --- | --- | --- | --- | --- | --- | --- |
|  | Testing |  | Positivity |  | Testing |  | Positivity |  | Testing |  | Positivity |  |
|  | OR (95% CI) | P-value | OR (95% CI) | P-value | OR (95% CI) | P-value | OR (95% CI) | P-value | OR (95% CI) | P-value | OR (95% CI) | P-value |
| **Sex<sup>a</sup>** |  | <0.001 |  | <0.001 |  | <0.001 |  | <0.001 |  | <0.001 |  | <0.001 |
| Female | Reference |  | Reference |  | Reference |  | Reference |  | Reference |  | Reference |  |
| Male | 0.62 (0.62-0.63) |  | 1.09 (1.05-1.12) |  | 0.63 (0.63-0.63) |  | 3.28 (3.11-3.45) |  | 1.16 (1.15-1.17) |  | 5.18 (4.79-5.61) |  |
| Unknown | 0.66 (0.63-0.69) |  | 0.79 (0.57-1.09) |  | 0.65 (0.62-0.68) |  | 2.31 (1.51-3.53) |  | 1.05 (1.01-1.08) |  | 5.13 (3.6-7.32) |  |
| **Race or Ethnicity<sup>b</sup>** |  | <0.001 |  | <0.001 |  | <0.001 |  | <0.001 |  | <0.001 |  | <0.001 |
| Asian or Pacific Islander | 0.98 (0.97-0.99) |  | 1.00 (0.93-1.08) |  | 0.95 (0.94-0.95) |  | 0.50 (0.43-0.58) |  | 1.03 (1.02-1.04) |  | 0.33 (0.26-0.42) |  |
| Black or African American | 1.06 (1.05-1.07) |  | 3.42 (3.24-3.61) |  | 1.06 (1.05-1.07) |  | 4.08 (3.73-4.46) |  | 1.18 (1.17-1.19) |  | 3.18 (2.82-3.57) |  |
| Hispanic or Latino | 1.13 (1.12-1.14) |  | 2.32 (2.19-2.46) |  | 1.13 (1.12-1.14) |  | 1.34 (1.21-1.49) |  | 1.23 (1.22-1.24) |  | 1.74 (1.53-1.99) |  |
| White | Reference |  | Reference |  | Reference |  | Reference |  | Reference |  | Reference |  |
| Other race<sup>c</sup> | 1.01 (1.00-1.02) |  | 1.83 (1.72-1.96) |  | 0.99 (0.98-1.00) |  | 1.44 (1.28-1.61) |  | 1.03 (1.02-1.04) |  | 1.45 (1.25-1.68) |  |
| Unknown | 1.17 (1.16-1.18) |  | 1.86 (1.76-1.98) |  | 1.14 (1.13-1.15) |  | 1.34 (1.21-1.48) |  | 1.18 (1.17-1.19) |  | 1.36 (1.19-1.54) |  |
| **Age Group** |  | <0.001 |  | <0.001 |  | <0.001 |  | <0.001 |  | <0.001 |  | <0.001 |
| 18-19 years | Reference |  | Reference |  | Reference |  | Reference |  | Reference |  | Reference |  |
| 20-24 years | 0.55 (0.53-0.56) |  | 0.70 (0.66-0.74) |  | 0.50 (0.49-0.52) |  | 0.75 (0.67-0.83) |  | 1.04 (1.02-1.07) |  | 2.05 (1.48-2.84) |  |
| 25-29 years | 0.63 (0.62-0.64) |  | 0.38 (0.35-0.40) |  | 0.58 (0.57-0.59) |  | 0.54 (0.49-0.60) |  | 1.11 (1.09-1.14) |  | 3.33 (2.43-4.57) |  |
| 30-34 years | 0.45 (0.44-0.45) |  | 0.20 (0.19-0.21) |  | 0.41 (0.40-0.42) |  | 0.36 (0.32-0.40) |  | 1.10 (1.07-1.12) |  | 3.94 (2.88-5.39) |  |
| 35-39 years | 0.52 (0.51-0.53) |  | 0.12 (0.11-0.14) |  | 0.47 (0.47-0.48) |  | 0.25 (0.22-0.29) |  | 1.05 (1.02-1.07) |  | 3.49 (2.54-4.78) |  |
| 40-44 years | 0.36 (0.35-0.37) |  | 0.08 (0.07-0.09) |  | 0.33 (0.32-0.33) |  | 0.17 (0.15-0.20) |  | 0.86 (0.84-0.88) |  | 2.81 (2.03-3.88) |  |
| 45-49 years | 0.40 (0.39-0.41) |  | 0.06 (0.05-0.06) |  | 0.37 (0.36-0.37) |  | 0.12 (0.10-0.14) |  | 0.78 (0.76-0.79) |  | 3.09 (2.23-4.27) |  |
| 50-54 years | 0.27 (0.26-0.27) |  | 0.04 (0.04-0.05) |  | 0.24 (0.24-0.25) |  | 0.07 (0.06-0.09) |  | 0.67 (0.65-0.69) |  | 3.19 (2.31-4.40) |  |
| 55-59 years | 0.27 (0.27-0.28) |  | 0.03 (0.02-0.03) |  | 0.24 (0.24-0.25) |  | 0.06 (0.05-0.08) |  | 0.65 (0.63-0.66) |  | 3.24 (2.35-4.48) |  |
| 60-64 years | 0.19 (0.19-0.20) |  | 0.01 (0.01-0.02) |  | 0.17 (0.17-0.17) |  | 0.02 (0.02-0.03) |  | 0.53 (0.52-0.54) |  | 2.29 (1.64-3.18) |  |
| 65+ years | 0.08 (0.08-0.08) |  | 0.005 (0.004-0.006) |  | 0.07 (0.06-0.07) |  | 0.01 (0.01-0.01) |  | 0.39 (0.38-0.40) |  | 1.07 (0.77-1.50) |  |
| **Borough of Residence** |  | <0.001 |  | <0.001 |  | <0.001 |  | <0.001 |  | <0.001 |  | <0.001 |
| Bronx | Reference |  | Reference |  | Reference |  | Reference |  | Reference |  | Reference |  |
| Brooklyn | 1.02 (1.01-1.03) |  | 0.87 (0.83-0.91) |  | 1.04 (1.03-1.05) |  | 0.88 (0.81-0.95) |  | 1.04 (1.03-1.05) |  | 0.67 (0.61-0.74) |  |
| Manhattan | 0.92 (0.91-0.93) |  | 0.92 (0.87-0.96) |  | 0.94 (0.93-0.95) |  | 1.19 (1.09-1.29) |  | 0.94 (0.93-0.95) |  | 0.83 (0.74-0.92) |  |
| Queens | 0.93 (0.92-0.94) |  | 0.98 (0.93-1.03) |  | 0.97 (0.96-0.98) |  | 0.80 (0.72-0.88) |  | 0.95 (0.94-0.96) |  | 0.50 (0.45-0.56) |  |
| Staten Island | 0.90 (0.89-0.92) |  | 1.00 (0.93-1.09) |  | 0.96 (0.95-0.98) |  | 1.26 (1.10-1.44) |  | 0.78 (0.77-0.79) |  | 0.55 (0.44-0.68) |  |
| **Area-Based Poverty Level<sup>d</sup>** |  | <0.001 |  | <0.001 |  | <0.001 |  | <0.001 |  | <0.001 |  | <0.001 |
| Low (<10% below FPT) | Reference |  | Reference |  | Reference |  | Reference |  | Reference |  | Reference |  |
| Medium (10% to <20%) | 0.98 (0.97-0.98) |  | 1.12 (1.07-1.17) |  | 0.93 (0.93-0.94) |  | 1.31 (1.20-1.43) |  | 0.99 (0.99-1.00) |  | 1.56 (1.38-1.76) |  |
| High (20% to <30%) | 1.00 (0.99-1.01) |  | 1.36 (1.29-1.43) |  | 0.97 (0.96-0.98) |  | 1.61 (1.45-1.77) |  | 1.00 (0.99-1.01) |  | 1.49 (1.30-1.71) |  |
| Very high (30% to 100%) | 0.94 (0.93-0.95) |  | 1.41 (1.33-1.50) |  | 0.90 (0.89-0.91) |  | 1.91 (1.72-2.12) |  | 1.01 (1.00-1.02) |  | 1.79 (1.55-2.07) |  |
| Unknown | 1.07 (0.97-1.18) |  | 1.29 (0.71-2.35) |  | 1.04 (0.94-1.15) |  | 1.16 (0.51-2.64) |  | 1.02 (0.93-1.12) |  | 3.13 (1.39-7.06) |  |

Abbreviations: ABPL, Area-based Poverty Level. CI, Confidence Interval. FPT, federal poverty threshold. HIV, human immunodeficiency virus. OMOP, Observational Medical Outcomes Partnership. OR, Odds Ratio. CDM, Common Data Model. <sup>a</sup>The OMOP CDM uses the term "gender" while source data were based on sex; source values may represent sex assigned at birth and/or administrative sex used for billing purposes (which may reflect gender identity, not sex assigned at birth). <sup>b</sup>All categories other than "Hispanic or Latino" are exclusively non-Hispanic or Latino (e.g., non-Hispanic White). <sup>c</sup>"Other Race" category is derived directly from reporting of "Other", "Other race", or similar in the source data. <sup>d</sup>ABPL categorized as low (<10% of residents in ZIP code tabulation area living below the FPT), medium (10% to <20%), high (20 to <30%), and very high (30 to 100%).

**Table S3.** Unadjusted odds ratios for laboratory testing and positive test results by sexually transmitted infection and patient characteristics.

| Characteristics | Chlamydia |  |  |  | Gonorrhea |  |  |  | HIV |  |  |  |
| --- | --- | --- | --- | --- | --- | --- | --- | --- | --- | --- | --- | --- |
|  | Testing |  | Positivity |  | Testing |  | Positivity |  | Testing |  | Positivity |  |
|  | OR (95% CI) | P-value | OR (95% CI) | P-value | OR (95% CI) | P-value | OR (95% CI) | P-value | OR (95% CI) | P-value | OR (95% CI) | P-value |
| **Sex<sup>a</sup>** |  | <0.001 |  | <0.001 |  | <0.001 |  | <0.001 |  | <0.001 |  | <0.001 |
| Female | Reference |  | Reference |  | Reference |  | Reference |  | Reference |  | Reference |  |
| Male | 0.59 (0.59-0.60) |  | 0.94 (0.91-0.97) |  | 0.60 (0.60-0.60) |  | 2.85 (2.70-3.01) |  | 1.11 (1.10-1.11) |  | 4.96 (4.59-5.36) |  |
| Unknown | 0.33 (0.31-0.34) |  | 0.25 (0.18-0.35) |  | 0.31 (0.30-0.32) |  | 0.74 (0.47-1.15) |  | 0.69 (0.67-0.71) |  | 3.26 (2.30-4.62) |  |
| **Race or Ethnicity<sup>b</sup>** |  | <0.001 |  | <0.001 |  | <0.001 |  | <0.001 |  | <0.001 |  | <0.001 |
| Asian or Pacific Islander | 1.00 (0.99-1.01) |  | 1.01 (0.94-1.09) |  | 0.97 (0.96-0.98) |  | 0.52 (0.44-0.61) |  | 1.07 (1.06-1.08) |  | 0.34 (0.27-0.43) |  |
| Black or African American | 1.17 (1.16-1.18) |  | 4.30 (4.07-4.54) |  | 1.16 (1.15-1.17) |  | 5.30 (4.86-5.79) |  | 1.27 (1.26-1.28) |  | 3.80 (3.39-4.25) |  |
| Hispanic or Latino | 1.33 (1.32-1.34) |  | 3.42 (3.23-3.62) |  | 1.33 (1.32-1.34) |  | 1.92 (1.74-2.12) |  | 1.36 (1.35-1.38) |  | 2.11 (1.87-2.38) |  |
| White | Reference |  | Reference |  | Reference |  | Reference |  | Reference |  | Reference |  |
| Other race<sup>c</sup> | 1.14 (1.13-1.15) |  | 2.42 (2.27-2.58) |  | 1.11 (1.10-1.13) |  | 1.94 (1.73-2.17) |  | 1.22 (1.20-1.23) |  | 1.71 (1.48-1.97) |  |
| Unknown | 1.29 (1.27-1.30) |  | 2.34 (2.21-2.48) |  | 1.25 (1.24-1.26) |  | 1.75 (1.58-1.94) |  | 1.18 (1.17-1.19) |  | 1.62 (1.42-1.84) |  |
| **Age Group** |  | <0.001 |  | <0.001 |  | <0.001 |  | <0.001 |  | <0.001 |  | <0.001 |
| 18-19 years | Reference |  | Reference |  | Reference |  | Reference |  | Reference |  | Reference |  |
| 20-24 years | 0.52 (0.51-0.54) |  | 0.67 (0.63-0.71) |  | 0.47 (0.46-0.48) |  | 0.69 (0.62-0.78) |  | 1.03 (1.01-1.06) |  | 1.82 (1.31-2.52) |  |
| 25-29 years | 0.64 (0.63-0.66) |  | 0.35 (0.33-0.37) |  | 0.59 (0.57-0.60) |  | 0.49 (0.43-0.55) |  | 1.10 (1.07-1.12) |  | 2.94 (2.14-4.03) |  |
| 30-34 years | 0.42 (0.41-0.43) |  | 0.18 (0.17-0.19) |  | 0.38 (0.37-0.39) |  | 0.32 (0.28-0.36) |  | 1.08 (1.06-1.11) |  | 3.53 (2.58-4.82) |  |
| 35-39 years | 0.52 (0.51-0.53) |  | 0.11 (0.10-0.12) |  | 0.47 (0.46-0.48) |  | 0.23 (0.19-0.26) |  | 1.03 (1.01-1.06) |  | 3.15 (2.30-4.33) |  |
| 40-44 years | 0.34 (0.33-0.35) |  | 0.08 (0.07-0.08) |  | 0.30 (0.29-0.31) |  | 0.16 (0.14-0.19) |  | 0.86 (0.84-0.88) |  | 2.63 (1.90-3.63) |  |
| 45-49 years | 0.39 (0.38-0.40) |  | 0.05 (0.04-0.06) |  | 0.35 (0.34-0.36) |  | 0.11 (0.09-0.13) |  | 0.77 (0.75-0.79) |  | 2.97 (2.15-4.10) |  |
| 50-54 years | 0.25 (0.25-0.26) |  | 0.04 (0.03-0.05) |  | 0.22 (0.22-0.23) |  | 0.07 (0.06-0.09) |  | 0.67 (0.65-0.68) |  | 3.26 (2.37-4.50) |  |
| 55-59 years | 0.26 (0.25-0.26) |  | 0.03 (0.02-0.03) |  | 0.22 (0.22-0.23) |  | 0.07 (0.05-0.09) |  | 0.64 (0.63-0.66) |  | 3.43 (2.49-4.73) |  |
| 60-64 years | 0.18 (0.18-0.18) |  | 0.01 (0.01-0.02) |  | 0.16 (0.15-0.16) |  | 0.02 (0.02-0.04) |  | 0.52 (0.51-0.54) |  | 2.33 (1.67-3.24) |  |
| 65+ years | 0.07 (0.07-0.08) |  | 0.004 (0.003-0.005) |  | 0.06 (0.06-0.06) |  | 0.01 (0.01-0.01) |  | 0.38 (0.37-0.39) |  | 0.98 (0.70-1.36) |  |
| **Borough of Residence** |  | <0.001 |  | <0.001 |  | <0.001 |  | <0.001 |  | <0.001 |  | <0.001 |
| Bronx | Reference |  | Reference |  | Reference |  | Reference |  | Reference |  | Reference |  |
| Brooklyn | 0.93 (0.92-0.94) |  | 0.63 (0.60-0.65) |  | 0.94 (0.94-0.95) |  | 0.67 (0.62-0.72) |  | 0.95 (0.95-0.96) |  | 0.54 (0.49-0.59) |  |
| Manhattan | 0.85 (0.84-0.86) |  | 0.58 (0.56-0.61) |  | 0.87 (0.87-0.88) |  | 0.72 (0.66-0.78) |  | 0.86 (0.85-0.86) |  | 0.61 (0.55-0.67) |  |
| Queens | 0.82 (0.81-0.83) |  | 0.54 (0.52-0.57) |  | 0.85 (0.84-0.86) |  | 0.39 (0.36-0.42) |  | 0.85 (0.85-0.86) |  | 0.32 (0.29-0.36) |  |
| Staten Island | 0.80 (0.78-0.81) |  | 0.52 (0.48-0.56) |  | 0.85 (0.84-0.86) |  | 0.60 (0.52-0.68) |  | 0.68 (0.67-0.69) |  | 0.31 (0.26-0.38) |  |
| **Area-Based Poverty Level<sup>d</sup>** |  | <0.001 |  | <0.001 |  | <0.001 |  | <0.001 |  | <0.001 |  | <0.001 |
| Low (<10% below FPT) | Reference |  | Reference |  | Reference |  | Reference |  | Reference |  | Reference |  |
| Medium (10% to <20%) | 1.06 (1.05-1.07) |  | 1.32 (1.26-1.38) |  | 1.01 (1.00-1.02) |  | 1.51 (1.38-1.65) |  | 1.09 (1.08-1.09) |  | 1.91 (1.69-2.15) |  |
| High (20% to <30%) | 1.14 (1.13-1.15) |  | 1.77 (1.69-1.86) |  | 1.09 (1.09-1.10) |  | 2.28 (2.08-2.51) |  | 1.15 (1.14-1.16) |  | 2.30 (2.02-2.61) |  |
| Very high (30% to 100%) | 1.16 (1.15-1.17) |  | 2.30 (2.19-2.42) |  | 1.10 (1.09-1.11) |  | 3.30 (3.01-3.62) |  | 1.23 (1.22-1.24) |  | 3.71 (3.28-4.21) |  |
| Unknown | 1.12 (1.02-1.23) |  | 1.44 (0.79-2.63) |  | 1.08 (0.98-1.19) |  | 1.32 (0.56-3.14) |  | 1.10 (1.00-1.20) |  | 4.93 (2.20-11.10) |  |

Abbreviations: ABPL, Area-based Poverty Level. CI, Confidence Interval. FPT, federal poverty threshold. HIV, human immunodeficiency virus. OMOP, Observational Medical Outcomes Partnership. OR, Odds Ratio. CDM, Common Data Model. <sup>a</sup>The OMOP CDM uses the term "gender" while source data were based on sex; source values may represent sex assigned at birth and/or administrative sex used for billing purposes (which may reflect gender identity, not sex assigned at birth). <sup>b</sup>All categories other than "Hispanic or Latino" are exclusively non-Hispanic or Latino (e.g., non-Hispanic White). <sup>c</sup>"Other Race" category is derived directly from reporting of "Other", "Other race", or similar in the source data. <sup>d</sup>ABPL categorized as low (<10% of residents in ZIP code tabulation area living below the FPT), medium (10% to <20%), high (20 to <30%), and very high (30 to 100%).

**Table S4.** Neighborhood-level spatial autocorrelation of laboratory testing and positive test results.

| STI | Testing |  |  | Positive Cases |  |  |
| --- | --- | --- | --- | --- | --- | --- |
|  | Moran's I | z-score | P-value | Moran's I | z-score | P-value |
| Chlamydia | 0.21 | 2.16 | 0.02 | 0.20 | 2.06 | 0.02 |
| Gonorrhea | 0.21 | 2.11 | 0.02 | 0.18 | 1.84 | 0.03 |
| HIV | 0.24 | 2.39 | 0.01 | 0.23 | 2.30 | 0.01 |

*Abbreviations:* HIV, human immunodeficiency virus. STI, sexually transmitted infection.

**Table S5.** Citywide percentages of laboratory tests and positive results by neighborhood.

| Borough | UHF Code and Neighborhood Name | Chlamydia |  |  | Gonorrhea |  |  | HIV |  |  |
| --- | --- | --- | --- | --- | --- | --- | --- | --- | --- | --- |
|  |  | Cases | Tests | Absolute Difference | Cases | Tests | Absolute Difference | Cases | Tests | Absolute Difference |
|  |  | % |  |  |  |  |  |  |  |  |
| Bronx | 101 Kingsbridge - Riverdale | 0.7 | 0.7 | 0 | 0.6 | 0.7 | -0.1 | 0.6 | 0.8 | -0.2 |
| Bronx | 102 Northeast Bronx | 2 | 1.4 | 0.6 | 2.1 | 1.4 | 0.7 | 2.1 | 1.4 | 0.7 |
| Bronx | 103 Fordham - Bronx Park | 3.8 | 3.1 | 0.7 | 2.8 | 3 | -0.2 | 5.8 | 2.8 | 3 |
| Bronx | 104 Pelham - Throgs Neck | 3.3 | 3.2 | 0.1 | 2.5 | 3.1 | -0.6 | 2.9 | 3 | -0.1 |
| Bronx | 105 Crotona - Tremont | 4.8 | 3.8 | 1 | 5.1 | 3.6 | 1.5 | 6.1 | 3.2 | 2.9 |
| Bronx | 106 High Bridge - Morrisania | 4.3 | 3.1 | 1.2 | 4.7 | 3 | 1.7 | 5 | 2.9 | 2.1 |
| Bronx | 107 Hunts Point - Mott Haven | 3.8 | 2.6 | 1.2 | 3.8 | 2.5 | 1.3 | 4.5 | 2.1 | 2.4 |
| Brooklyn | 201 Greenpoint | 1 | 1.6 | -0.6 | 1.5 | 1.6 | -0.1 | 0.7 | 1.6 | -0.9 |
| Brooklyn | 202 Downtown - Heights - Park Slope | 1.6 | 2.4 | -0.8 | 2.4 | 2.5 | -0.1 | 2.1 | 2.7 | -0.6 |
| Brooklyn | 203 Bedford Stuyvesant - Crown Heights | 5.9 | 4.5 | 1.4 | 7.6 | 4.6 | 3 | 7.8 | 4.7 | 3.1 |
| Brooklyn | 204 East New York | 2.1 | 1.4 | 0.7 | 2 | 1.4 | 0.6 | 1.8 | 1.3 | 0.5 |
| Brooklyn | 205 Sunset Park | 4.2 | 4.2 | 0 | 3.1 | 4.2 | -1.1 | 3.2 | 4.3 | -1.1 |
| Brooklyn | 206 Borough Park | 1.9 | 3.3 | -1.4 | 1.2 | 3.3 | -2.1 | 1.7 | 3.8 | -2.1 |
| Brooklyn | 207 East Flatbush - Flatbush | 4.3 | 3.6 | 0.7 | 5.1 | 3.6 | 1.5 | 6.3 | 3.6 | 2.7 |
| Brooklyn | 208 Canarsie - Flatlands | 2.3 | 2 | 0.3 | 1.8 | 2 | -0.2 | 2 | 2.3 | -0.3 |
| Brooklyn | 209 Bensonhurst - Bay Ridge | 1.2 | 2.3 | -1.1 | 0.7 | 2.3 | -1.6 | 1.2 | 2.6 | -1.4 |
| Brooklyn | 210 Coney Island - Sheepshead Bay | 1.9 | 3.1 | -1.2 | 1.6 | 2.9 | -1.3 | 1.7 | 3 | -1.3 |
| Brooklyn | 211 Williamsburg - Bushwick | 4 | 3.5 | 0.5 | 4.9 | 3.5 | 1.4 | 2.8 | 3 | -0.2 |
| Manhattan | 301 Washington Heights - Inwood | 4.2 | 3.5 | 0.7 | 4 | 3.4 | 0.6 | 3.8 | 3.3 | 0.5 |
| Manhattan | 302 Central Harlem - Morningside Heights/ Manhattan | 3.1 | 2.5 | 0.6 | 5.5 | 2.5 | 3 | 3.8 | 2.3 | 1.5 |
| Manhattan | 303 East Harlem | 2.6 | 1.8 | 0.8 | 3 | 1.7 | 1.3 | 2.5 | 1.9 | 0.6 |
| Manhattan | 304 Upper West Side | 1 | 1.5 | -0.5 | 1.2 | 1.5 | -0.3 | 1.7 | 1.7 | 0 |
| Manhattan | 305 Upper East Side | 1 | 1.8 | -0.8 | 1.1 | 1.9 | -0.8 | 0.9 | 1.9 | -1 |
| Manhattan | 306 Chelsea - Clinton | 1.8 | 2.4 | -0.6 | 4.1 | 2.4 | 1.7 | 4.4 | 2.2 | 2.2 |
| Manhattan | 307 Gramercy Park - Murray Hill | 1 | 1.5 | -0.5 | 1.1 | 1.5 | -0.4 | 1.2 | 1.4 | -0.2 |
| Manhattan | 308 Greenwich Village - SoHo | 0.4 | 0.8 | -0.4 | 0.5 | 0.8 | -0.3 | 0.3 | 0.7 | -0.4 |
| Manhattan | 309 Union Square - Lower East Side | 1.7 | 1.9 | -0.2 | 1.8 | 1.9 | -0.1 | 1.2 | 2 | -0.8 |
| Manhattan | 310 Lower Manhattan | 0.6 | 0.7 | -0.1 | 0.7 | 0.7 | 0 | 0.5 | 0.7 | -0.2 |
| Queens | 401 Long Island City - Astoria | 2.3 | 2.9 | -0.6 | 2.3 | 2.9 | -0.6 | 2 | 3 | -1 |
| Queens | 402 West Queens | 5.1 | 5.9 | -0.8 | 3.5 | 5.8 | -2.3 | 3.9 | 6 | -2.1 |
| Queens | 403 Flushing - Clearview | 2.4 | 3.4 | -1 | 1.4 | 3.4 | -2 | 0.9 | 3.3 | -2.4 |
| Queens | 404 Bayside - Little Neck | 0.5 | 0.8 | -0.3 | 0.3 | 0.8 | -0.5 | 0.3 | 0.9 | -0.6 |
| Queens | 405 Ridgewood - Forest Hills | 1.8 | 2.7 | -0.9 | 1 | 2.8 | -1.8 | 1.2 | 2.4 | -1.2 |
| Queens | 406 Fresh Meadows | 0.9 | 1.1 | -0.2 | 0.6 | 1.1 | -0.5 | 0.2 | 1.2 | -1 |
| Queens | 407 Southwest Queens | 3 | 3.2 | -0.2 | 1.8 | 3.2 | -1.4 | 2.2 | 3.5 | -1.3 |
| Queens | 408 Jamaica | 5.2 | 4.1 | 1.1 | 4 | 4.2 | -0.2 | 4.4 | 4.5 | -0.1 |
| Queens | 409 Southeast Queens | 2.8 | 2.3 | 0.5 | 2.3 | 2.4 | -0.1 | 1.6 | 2.6 | -1 |
| Queens | 410 Rockaway | 1.6 | 1.2 | 0.4 | 1.6 | 1.2 | 0.4 | 1.4 | 1.3 | 0.1 |
| Staten Island | 501 Port Richmond | 1 | 0.8 | 0.2 | 1 | 0.8 | 0.2 | 0.6 | 0.7 | -0.1 |
| Staten Island | 502 Stapleton - St. George | 1.7 | 1.4 | 0.3 | 2.3 | 1.4 | 0.9 | 1.6 | 1.5 | 0.1 |
| Staten Island | 503 Willowbrook | 0.6 | 0.7 | -0.1 | 0.5 | 0.7 | -0.2 | 0.4 | 0.7 | -0.3 |
| Staten Island | 504 South Beach - Tottenville | 1 | 1.6 | -0.6 | 0.9 | 1.7 | -0.8 | 0.5 | 1.3 | -0.8 |

Abbreviations: UHF, United Hospital Fund. Case percentages determined by the number of positive cases identified in a given neighborhood divided by the total number of positive cases found in all of New York City. Test percentages determined by the number of laboratory tests performed in a given neighborhood divided by the total number of tests performed in all of New York City. Columns labeled "Cases" and "Tests" may not total to 100% due to rounding. Absolute difference calculated by subtracting the percent of tests conducted in a given neighborhood from the percent of positive cases identified in that same neighborhood.

